# Early Prediction of In-Hospital Death of COVID-19 Patients: A Machine-Learning Model Based on Age, Blood Analyses, and Chest X-Ray Score

**DOI:** 10.1101/2021.06.10.21258721

**Authors:** E. Garrafa, M. Vezzoli, M. Ravanelli, D. Farina, A. Borghesi, S. Calza, R. Maroldi

## Abstract

**Background:** To develop and validate an early-warning model to predict in-hospital mortality on admission of COVID-19 patients at an emergency department (ED).

**Methods:** In total, 2782 patients were enrolled between March 2020 and December 2020, including 2106 patients (first wave) and 676 patients (second wave) in the COVID-19 outbreak in Italy. The first-wave patients were divided into two groups with 1474 patients used to train the model, and 632 to validate it. The 676 patients in the second wave were used to test the model. Age, 17 blood analytes and Brescia chest X-ray score were the variables processed using a Random Forests classification algorithm to build and validate the model. ROC analysis was used to assess the model performances. A web-based death-risk calculator was implemented and integrated within the Laboratory Information System of the hospital.

**Results:** The final score was constructed by age (the most powerful predictor), blood analytes (the strongest predictors were lactate dehydrogenase, D-dimer, Neutrophil/Lymphocyte ratio, C-reactive protein, Lymphocyte %, Ferritin std and Monocyte %), and Brescia chest X-ray score. The areas under the receiver operating characteristic curve obtained for the three groups (training, validating and testing) were 0.98, 0.83 and 0.78, respectively.

**Conclusions:** The model predicts in-hospital mortality on the basis of data that can be obtained in a short time, directly at the ED on admission. It functions as a web-based calculator, providing a risk score which is easy to interpret. It can be used in the triage process to support the decision on patient allocation.

## Introduction

Starting from late February 2020, the COVID-19 outbreak struck the north of Italy causing more than 30,000 deaths in Lombardy alone, up to the end of March 2021. At the beginning of the outbreak, the Spedali Civili di Brescia (SCBH), the university hospital of one of the hardest hit cities in Europe, was faced with a ‘flash flood’ of severely ill patients seeking admission to the Emergency Department (ED). For several weeks, their number exceeded the available resources, obliging a continuous organizational restructuring of the hospital wards (Garrafa et al., 2020b).

In those weeks, given the limited evidence of clinically proven predictors (Marengoni et al., 2021)(Wynants et al., 2020)(Sperrin et al., 2020), prioritizing hospital admission of non-critical patients was an arduous task. Essentially, the criteria were based on the presence of fever, respiratory symptoms and the level of blood oxygenation. A significant drawback of this approach was that patients referring to the ED with very similar clinical findings underwent inconsistent assessments. In this scenario, the availability of predictors would have been extremely beneficial, not only to triage patients, but also to monitor hospitalized patients and warn of exacerbation of the outbreaks.

Starting from March 2020, all patients referred to EDs underwent a chest X-ray at admission or within a few hours. With the purpose of grading pulmonary involvement and tracking changes objectively over time, a chest X-ray severity score was developed (Brescia X-ray score) (Borghesi and Maroldi, 2020)(Maroldi et al., 2020)(Borghesi et al., 2020a)(Borghesi et al., 2020b). The score was able to predict in-hospital mortality in 302 patients. In addition to the chest X-ray severity score, a dedicated blood sampling profile was included in the COVID-19 ED work-up (Garrafa et al., 2020a). Among its 17 blood analytes, the sampling profile encompassed hemachrome, inflammation biomarkers such as C reactive protein (CRP), lactate dehydrogenase (LDH) and Ferritin, and coagulation markers (Fibrinogen and D-dimer). Since that time, the medical literature began to encompass an increasing number of studies advocating the prognostic value of single or grouped blood parameters (Bonetti et al., 2020)(Borghi et al., 2020)(Avouac et al., 2021). All of these parameters were present in our COVID-19 sampling profile.

This study aims to develop and validate an Early-Warning Model (BS-EWM), predictive of in-hospital death, based on data that could easily be acquired on admission to the ED: age, simple blood biomarkers and chest X-ray. The model was constructed based on the analysis of a cohort of 2872 COVID-19 patients treated in a single reference center over a 10-month period.

This paper adheres to the TRIPOD checklist for predictive model development and validation (Collins et al., 2015).

The study was approved by the local ethics committee (NP 4000).

## Materials and Methods

The dataset contained 2782 COVID-19 symptomatic patients, hospitalized between March and December 2020 at SCBH after referring to the ED. In all patients, the following variables were retrieved from the SCBH database: age, sex, length of hospitalization, Brescia X-ray score (Borghesi and Maroldi, 2020), Alive/Dead, and 17 blood analytes acquired at admission (D-dimer, Fibrinogen, lactate dehydrogenase (LDH), Neutrophils, Lymphocytes, Neutrophil/Lymphocyte ratio (NLR), Lymphocytes %, Neutrophils %, C-reactive protein (CRP), white blood cell (WBC) count, Basophils, Basophils %, Eosinophils, Eosinophils %, Monocytes, Monocytes %, standardized Ferritin). Blood tests were acquired within 24 hours after admission to the hospital.

According to the two temporal peaks of incidence of the COVID-19 outbreak in Lombardy, the 2782 patients were divided into two groups: (*i*) March–April (MA) including 2106 patients admitted during the first wave; (*ii*) May–December (MD) including 676 patients in the second wave. Quantitative variables were described using Mean (SD), Median (IQR) and Range (min–max), while categorical variables were reported as counts and percentages. The comparisons between groups were performed using the Wilcoxon rank-sum test for quantitative variables and Fisher’s exact test for qualitative variables.

The relationships between the 17 analytes and the Brescia X-ray score were inspected using the Spearman correlation coefficient, *ρ*_*s*_, and visualizing results using a correlation plot (Dancelli et al., 2013)(Marziano et al., 2019) (Figure S1).

To estimate the BS-EWM, the outcome (Alive/Dead) was modeled using as covariates: (*i*) Brescia X-ray score, (*ii*) 17 analytes, (*iii*) age. Since most of the covariates analyzed were strongly correlated (multi-collinearity) (Figure S1) and their relationships with the outcome were non-linear, the BS-EWM was estimated using Random Forests (Breiman, 2001)(Carpita and Vezzoli, 2012), a non-parametric machine-learning method (Vezzoli, 2011)(Vezzoli et al., 2017). Moreover, the algorithm is able to manage missing values which are common in clinical studies. The “*on-the-fly-imputation*” algorithm (Hong and Lynn, 2020) imputes data when it grows the forest handling interactions and non-linearity in the dataset.

Since the prevalence rate of death in the two waves was different (20% in MA vs 12% in MD), a strategy to generalize results in unbalanced datasets was applied, adopting a rebalancing method able to improve the detection of patients with a high death-risk.

The EWM was developed using the 2106 patients in the first COVID-19 wave (MA 2020) when in-hospital death prevalence was 20%. Seventy percent of them (1474 patients) were used for training the model and the remainder (632 patients) for testing it. Patients were randomly assigned to the two subgroups, and further stratified according to the outcome (Alive/Dead). Consequently, both the training and testing subgroups included the same rate of deaths (20.09%) as the full sample (2106 patients). With such a “moderate” incidence of death, the dataset was statistically unbalanced. This limitation could have implied the development of a model yielding unsatisfactory results in predicting new observations for the minority class, i.e., patients with death as outcome. An approach to address this limitation is to oversample the minority class (deceased patients) and, subsequently, create the predictive model (BS-EWM). The Synthetic Minority Oversampling Technique (SMOTE) (Chawla et al., 2002) was chosen. The SMOTE function oversamples the minority class by using bootstrapping and *k-*nearest neighbor to synthetically create additional observations belonging to that class (Dead). The procedure is combined with under-sampling of the majority class (Alive). To determine the optimum number of *k*-groups into which to assign the dataset, a matrix containing the 17 analytes and the Brescia X-ray score was used to compute the hierarchical cluster analysis (Salvi et al., 2019)(Codenotti et al., 2016). By means of silhouette analysis, *k*=2 was determined as the optimal number of clusters into which to assign the dataset. Hence, a synthetic rebalanced dataset was obtained with an equal number of Living and Deceased patients (888+888). The rebalancing procedure enabled a risk score to be devised ranging from 0 to 1 with a threshold of 0.5 to separate non-severely affected from severely affected patients. Subsequently, the model was tested on the subgroup of 632 patients in the first wave excluding the training set. A further validation of the EWM was conducted on the 676 COVID-19 patients in the second wave (MD 2020).

The Relative Variable Importance Measure (rel VIM)(Carpita and Vezzoli, 2012),(Doglietto et al., 2020b) and the Partial Dependence Plots (PDP) (Friedman, 2001)(Doglietto et al., 2020a) were extracted from the model for a better understanding of the relationship between outcome and covariates.

The predictions extracted from the Random Forests classification were interpreted as in-hospital death probability conditional on the combination of the values of analytes, Brescia X-ray score and age in COVID-19 patients at admission to the ED.

The BS-EWM performance was evaluated by Area Under the Curve (AUC) of a Receiver Operating Characteristic (ROC) curve. The robustness of the model was compared to other models by running Gradient Boosting Machine (GBM, a machine-learning approach and competitor to Random Forests), and Logistic Regression, and computing the same metrics.

The BS-EWM score is available for use online (https://bdbiomed.shinyapps.io/covid19score). In the SCBH it is integrated within LIS (Laboratory Information System) returning the death-risk score directly from the medical report.

## Results

### Description of the sample

The entire sample analyzed in this paper contained 2782 COVID-19 patients (1010 female (36.3%) and 1772 male (63.7%)), admitted to the ED and hospitalized at SCBH from March to December 2020. During these 10 months, the pandemic had two temporal waves: March–April (2106 patients, 75.70% of the entire sample) and May–December (676 patients, 24.30% of the entire sample) (Table S1). The model was trained on a subsample extracted from the first wave (70%) and tested (*i*) on data not used to calibrate the model (remaining 30% from the first wave) and (*ii*) on data from the second wave.

The first-wave subsample contained 2106 COVID-19 patients hospitalized in March–April 2020 at SCBH: 744 females (35.3%), and 1362 males (64.7%) (Table 1). During that period, 423 patients died (20.09% of the total): 131 females (31%) and 292 males (69%). Their mean age±SD was 66.89±14.19: 67.93±15.40 for females and 66.32±13.45 for males (p-value=0.001). The mean age of deceased patients was 76.21±9.12, while for living patients, it was 64.55±14.27 (p-value < 0.001). Mean hospital stay was 13.58±11.58 days (from a minimum of 0 to a maximum of 140 days): 11.33±10.98 days for patients who died, 14.15±11.66 days for surviving patients (p-value < 0.001).

**Table 1:**
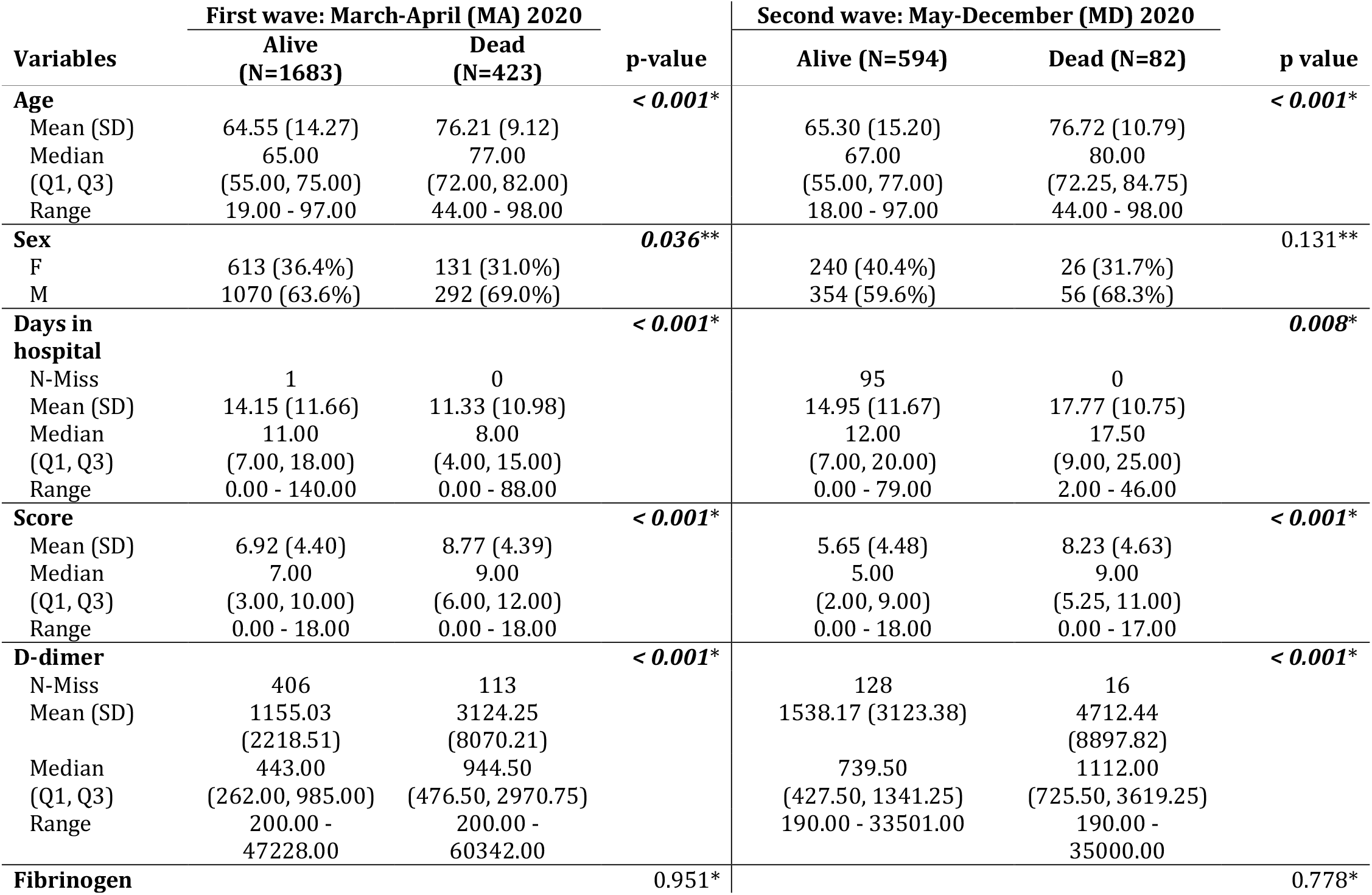
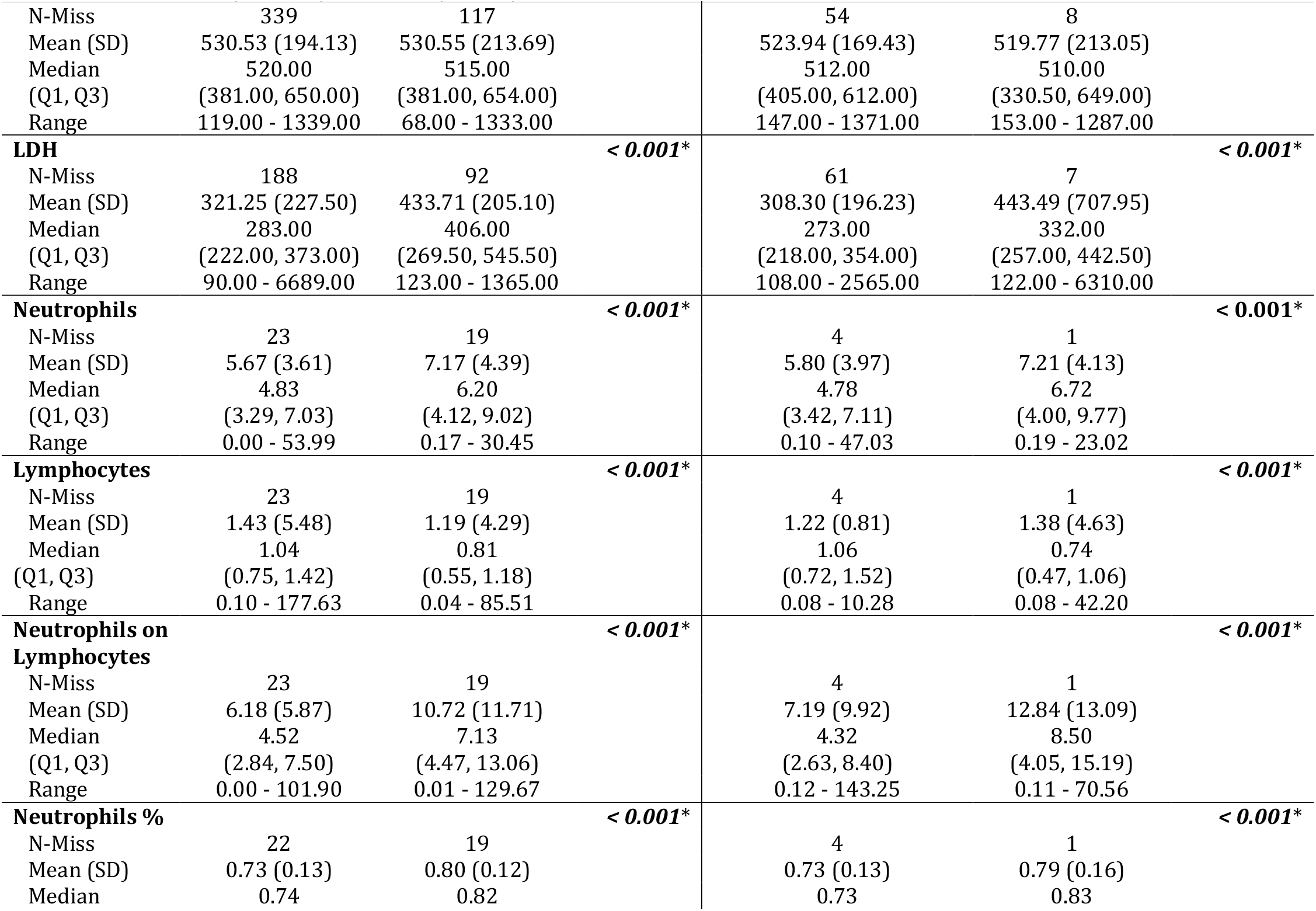
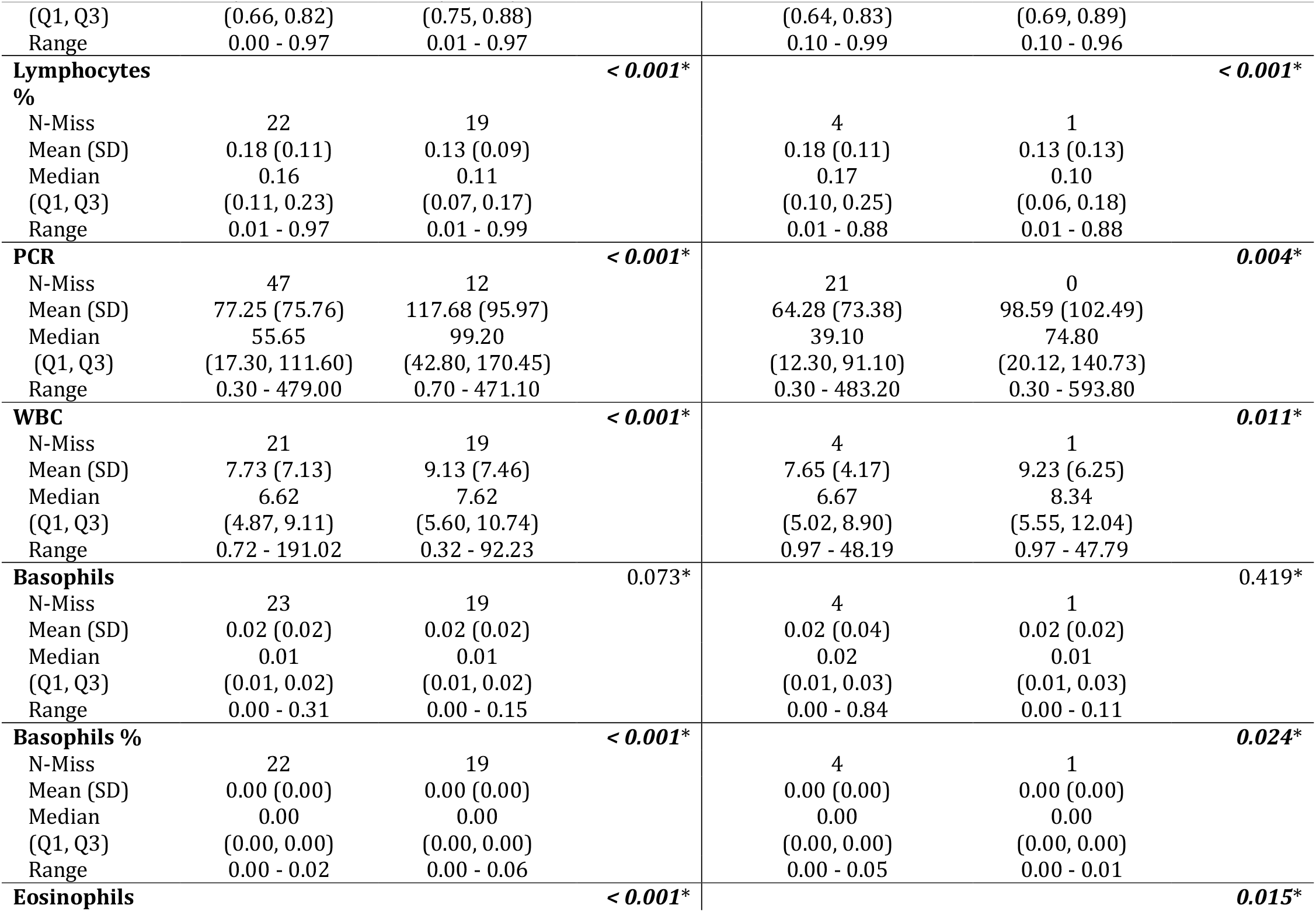
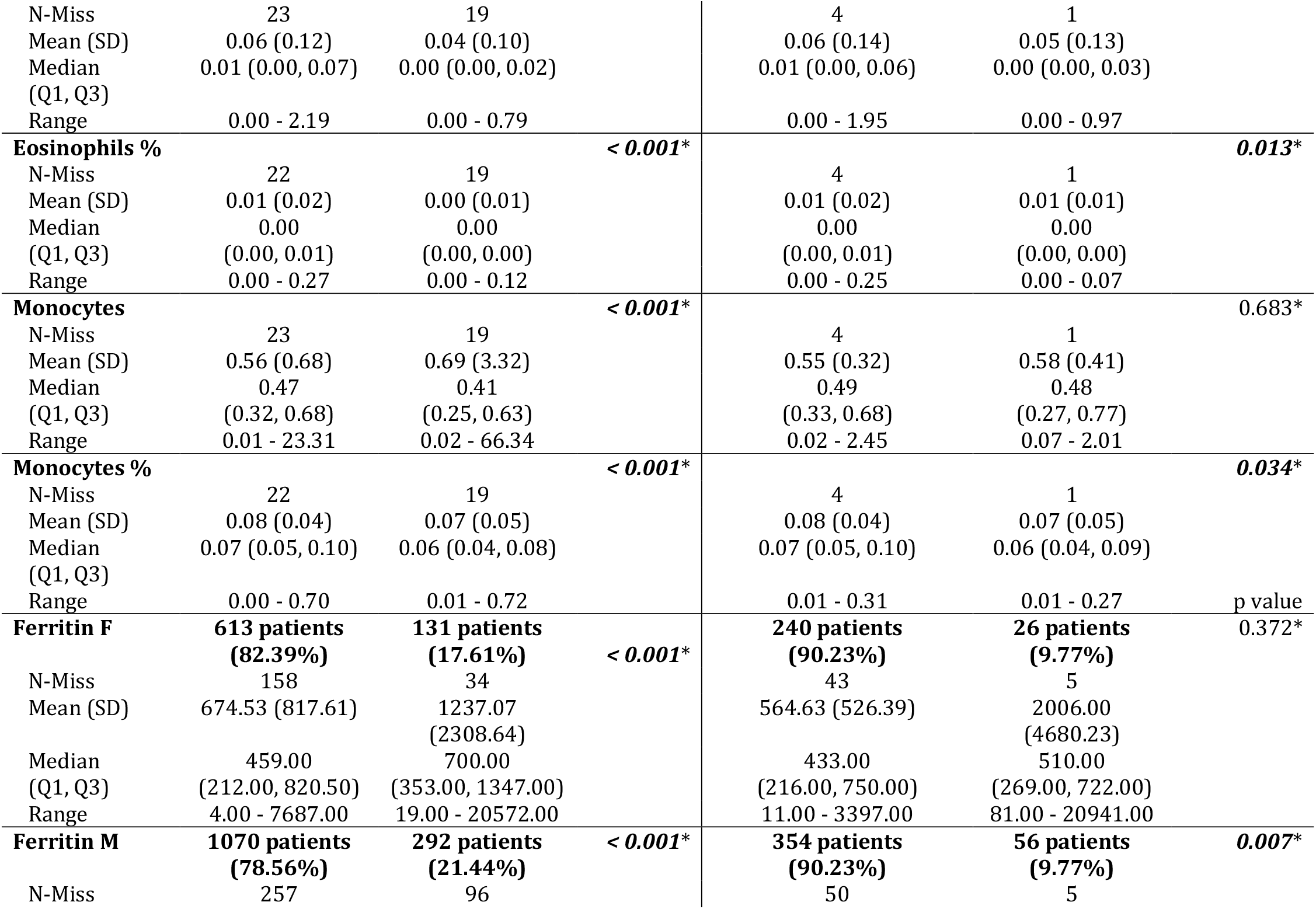
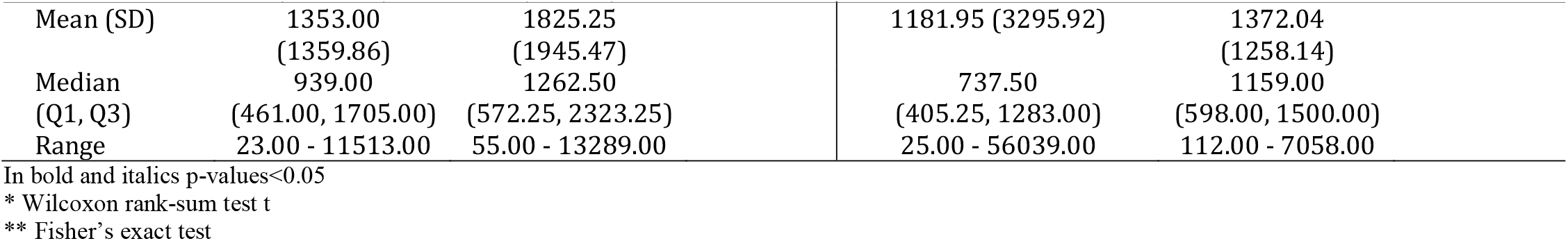
Descriptive statistics on all variables in the dataset stratified respect Alive-Dead. Comparison between first (March-April) and second (May-December) wave.

The second-wave subsample contained 676 COVID-19 patients hospitalized in May–December 2020 at SCBH: 266 females (39.3%), 410 males (60.7%) (Table 1). During the 8 months of the second wave, 82 patients died (12.13%): 26 females (31.7%) and 56 males (68.3%). The mean age of deceased patients was 76.72±10.79 versus 65.30±15.20 for surviving patients (p-value < 0.001). The mean hospital stay was 15.35±11.58 days (from a minimum of 0 to a maximum 79 days): 17.77±10.75 days for patients who died, 14.95 ±11.67 days for surviving patients (p-value=0.008).

The descriptive statistics for all variables in the dataset are presented in Table S2 and were computed and stratified by the two waves (MA vs MD) and by outcome (Alive vs Dead). The two subsets were similar for most variables.

The correlations between the 17 analytes and the Brescia X-ray score were investigated using Spearman correlation coefficients and visualized using a correlation plot (Figure S1). The Brescia X-ray score was positively correlated with Neutrophil to Lymphocyte ratio, CRP, LDH, standardized Ferritin, and D-Dimer, and was negatively correlated with Lymphocyte %, Monocyte %, and Basophil %.

### BS-EWM

A machine-learning model (BS-EWM) was developed by inputting a dataset of 2782 COVID-19 patients admitted to the ED and hospitalized at SCBH from March to December 2020. The majority of patients (2106/2782, 75.70%) belonged to the first wave (MA), the remaining fraction (676/2782, 24.30%) to the second wave (MD). As outcome, the machine-learning model had the condition Dead/Alive, and, as covariates: age, Brescia X-ray score and 17 blood sample analytes.

Figure 1 reports the flow chart that describes how data were divided for training, validation and testing the BS-EWM.

**Figure 1:**
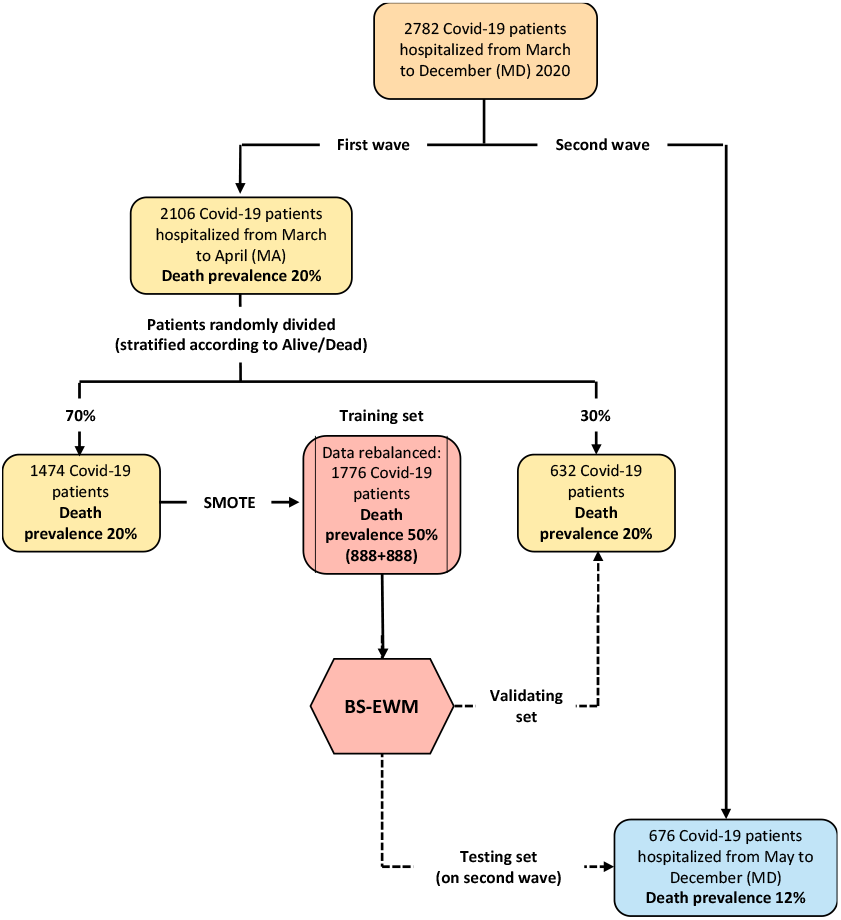
Flow-chart of the data used in the empirical analyses The BS-EWM was trained with a Random Forest on 70% of first wave patients (rebalanced with the SMOTE procedure) and (*i*) validated on remaining 30% of first wave patients (*ii*) tested on 676 second wave patients. In detail, 2106 patients were randomly in training and validating, maintaining the same death prevalence of the first wave.

The SMOTE procedure, rebalancing the Dead/Alive ratio (50% vs. 50%) from the original 20.09%, improved accuracy, specificity, and sensitivity of the Random Forest applied on it (see Table S3 which compares performance metrics with/without the SMOTE method).

The rel VIM and PDP were extracted from the Random Forests (Figure 2, panel A and B respectively). In panel A1, the rel VIM of BS-EWM based on age, Brescia X-ray score and 17 blood analytes are reported on a bar plot. Since age was strongly associated with the risk of death, it masked the role of the other covariates. For completeness, the relevance of the 17 analytes and Brescia X-ray score was estimated in an additional EWM, in which the covariate ‘age’ was excluded. In the resulting bar plot (Figure 2, panel A2), 9/17 analytes and the Brescia X-ray score were noted as being important in predicting the risk of death (rel VIM>60). The effects of changes in covariate values on the risk of death-threshold of the EWM were reported by means of a PDP (a 2D plot in the x–y plane) (Figure 2, panel B). Only Fibrinogen was excluded from this graphical representation since in Table 1, it was not significantly different in the two subpopulations Deceased/Alive. Most PDPs showed nonmonotonic increasing relationships between the *x*-variable and the EWM, resulting in a plateau corresponding to high values of *x*.

**Figure 2:**
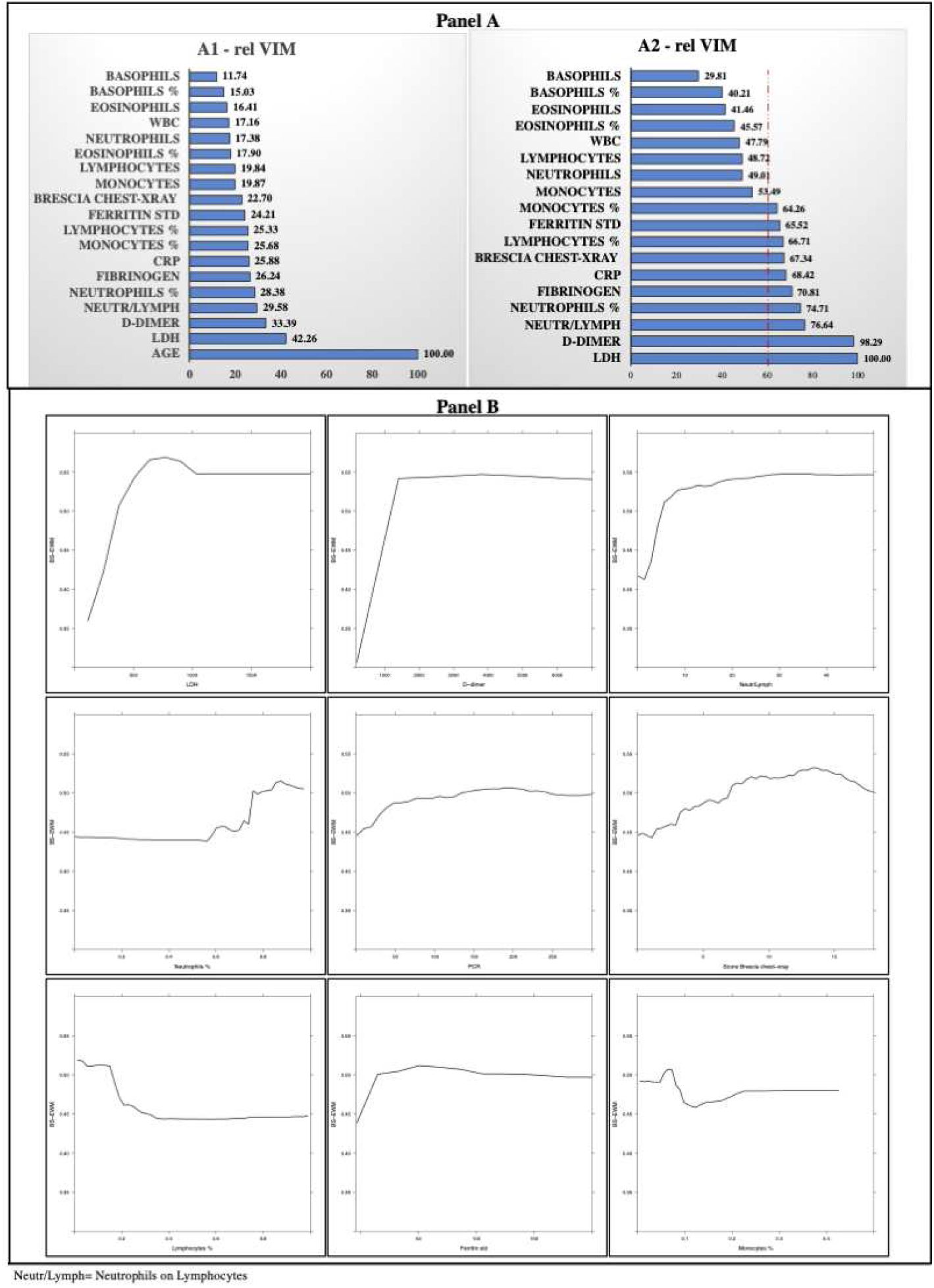
Relative Variable importance Measure (rel VIM) and PDP **PANEL A**: Relative Variable importance Measure (rel VIM) extracted from the Random Forest considering “Age” (A1) or excluding “Age” (A2). **Panel B**: Partial Dependence Plot (PDP) in correspondence of variables with VIM>60 (cut-off identified by the red dashed line) extracted from Random Forest without “Age” (A2) and p-value in Table 1 <0.05. PDPs are displayed from the most to the less important variable.

When compared to other models such as GBM and Logistic Regression, the Random Forest showed better performance in terms of AUC, sensitivity, and specificity. The in-sample sensitivity (0.93) yielded by the model was the highest, and it maintained an important 0.82 in validating the out-of-sample sensitivity, and this decreased to 0.73 when testing the MD subgroup (see Table 2 which contains details on all the metrics extracted from the ROC analysis). ROC curves are visualized in Figure 3 where, for each model (Random Forest, GBM and Logistic Regression), the performances in Training, Validating and Testing are compared in a unique graph.

**Table 2:** Performance metrics of methods: Random Forest, Gradient Boosting Machine (GBM) and Logistic Regression

| Metrics | Random Forest |  |  | GBM |  |  | LOGISTIC REGRESSION |  |  |
| --- | --- | --- | --- | --- | --- | --- | --- | --- | --- |
|  | Training<br>March-April (MA) | Validating<br>March-April (MA) | Testing<br>May-Dec<br>(MD) | Training<br>March-April (MA) | Validating<br>March-April (MA) | Testing<br>May-Dec<br>(MD) | Training<br>March-April (MA) | Validating<br>March-April (MA) | Testing<br>May-Dec<br>(MD) |
| **AUC**<br>**(DeLong)**<br>**(95% CI)** | 0.97<br>(0.97-0.98) | 0.83<br>(0.80-0.87) | 0.78<br>(0.73-0.84) | 0.88<br>(0.86-0.89) | 0.84<br>(0.80-0.88) | 0.78<br>(0.73-0.83) | 0.84<br>(0.82-0.86) | 0.83<br>(0.79-0.87) | 0.52<br>(0.44-0.60) |
| **Sensitivity**<br>**(95% CI)** | 0.93<br>(0.91-0.97) | 0.82<br>(0.72-0.92) | 0.73<br>(0.54-1.00) | 0.85<br>(0.80-0.88) | 0.80<br>(0.66-0.90) | 0.77<br>(0.65-0.94) | 0.80<br>(0.77-0.84) | 0.84<br>(0.76-0.91) | 0.87<br>(0.18-1.00) |
| **Specificity**<br>**(95% CI)** | 0.92<br>(0.88-0.94) | 0.75<br>(0.63-0.83) | 0.73<br>(0.41-0.89) | 0.77<br>(0.73-0.81) | 0.75<br>(0.65-0.87) | 0.71<br>(0.50-0.79) | 0.74<br>(0.70 -0.77) | 0.73<br>(0.65-0.79) | 0.26<br>(0.11-0.94) |

Comparison between the performances of three methods: Random Forest, GBM and Logistic Regression model applied on the rebalanced dataset obtained with SMOTE methodology. Logistic Regression predictions are computed using the 10-fold cross-validation in order to be comparable with Random Forest and GBM predictions (which use out-of-bag and 10-fold cross-validation, respectively).

**Figure 3:**
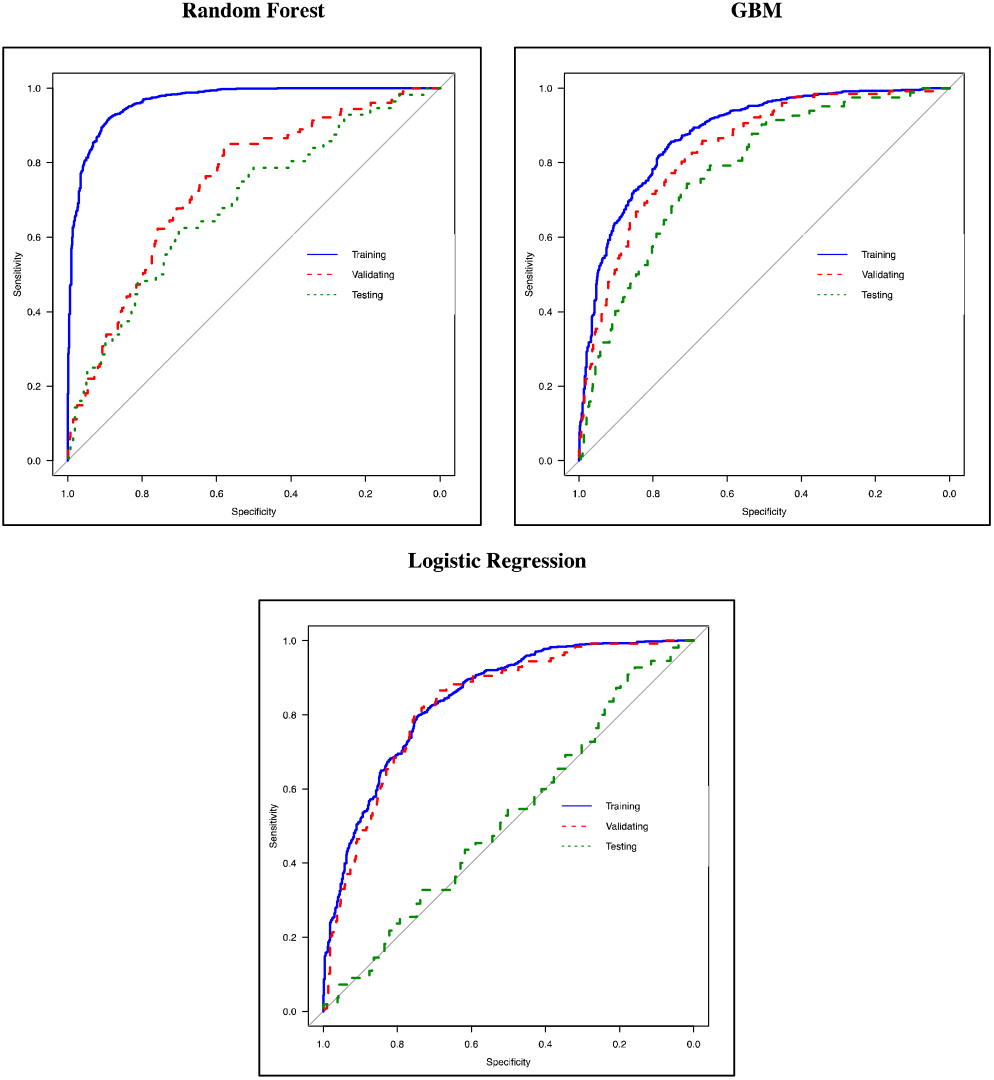
ROC curves of Random Forest, GBM and Logistic Regression ROC curves of three methods: (*i*) Random Forest, (*ii*) GBM and (*iii*) Logistic Regression. Each graph reports the ROC curve computed in Training (blue line, 70% of March-April’s patients) Validating (dashed red line, 30% of March-April’s patients), and Testing (dashed green lined, May-December patients).

## Discussion

The dataset for the development, validation and testing of the BS-EWM originated entirely from an Italian region, potentially limiting the generalizability of the risk score in other areas of the world. Additional validation studies from different geographic areas are welcomed. Furthermore, though the BS-EWM has been validated using blood sample values obtained by instruments that satisfy internal and external quality control, different equipment could lead to divergent results (Martens et al., 2021)(Lippi et al., 2020). Therefore, it would be appropriate to harmonize the results. Another limit could have been the presence of missing values, though the BS-EWM has also performed adequately in this condition since it used a multiple imputation technique to overcome the problem. Finally, it is important to point out that the BS-EWM risk score should not be used for asymptomatic COVID-19 patients or for the pediatric population.

Though the BS-EWM has been developed on a cohort of 2106 patients belonging to the COVID-19 first wave, the model also demonstrated a sensitivity greater than 70% in the early prediction of high risk in patients in the second wave, when in-hospital mortality was 40% lower.

Several predictive models have recently been applied to COVID-19 cohorts with variable results, some of them previously developed to predict mortality for community-acquired pneumonia, such as the Pneumonia Severity Index, CURB-65, qSOFA, and MuLBSTA(Yavuz et al., 2021)(Lazar Neto et al., 2021)(Artero et al., 2021), NEWS2 criteria (Myrstad et al., 2020)(Gidari et al., 2020), and SCAP score (Anurag and Preetam, n.d.). Novel early-warning scores have been specifically built on COVID-19 patient series using different techniques such as parametric and non-parametric tests (Linssen et al., 2020) or artificial intelligence techniques such as the COVID-GRAM score (Liang et al., 2020).

While these models are mostly based on age and a set of vital (clinical) parameters, in addition to age, the BS-EWM depends on blood parameters. It is conceivable that blood analytes capture a snapshot at hospital admission signaling a specific bodily reaction to viral infection in terms of hyperinflammation, immune response and thrombophilia. On the other hand, the other models are more influenced by the general status of the patient, which may be determined by concomitant and pre-existing diseases.

According to the International Federation of Clinical Chemistry (Bohn et al., 2020), no single biochemical or hematological marker is sufficiently sensitive or specific to predict the outcome of SARS-CoV-2 infection. Notably, the IFCC recommends that the interpretation of laboratory abnormalities should be based on groups of analytes (Bohn et al., 2020). In the BS-EWM, three analytes reached a significant value in predicting death: LDH, D-dimer and NLR. LDH is a non-specific indicator of tissue damage (Bohn et al., 2020)(Liang et al., 2020) that emerges as one of the most consistently elevated markers in patients at higher risk of developing adverse outcomes, probably because COVID-19 infection is characterized by systemic tissue damage. Another key feature of SARS-CoV-2 is the coagulopathy: high levels of D-dimers have been reported to correlate with unfavorable disease progression in several cohorts of patients. The coagulopathy linked to COVID-19 infection is likely to involve a complex interplay between pro-thrombotic and inflammatory factors, thus the combined analysis of both inflammatory and thrombophilic markers could play an important role in the early identification of patients at higher risk of unfavorable progression (Bohn et al., 2020)(Lazzaroni et al., 2021). Finally, lymphopenia has become a hallmark of SARS-CoV-2. It has been demonstrated in almost all symptomatic patients, though in varying degrees. Disease severity has been correlated with the level of lymphocyte count reduction. A direct infection of lymphocytes, which express the coronavirus receptor ACE-2, is among the mechanisms proposed. A poor prognosis is also associated with an elevated neutrophil count combined with lymphopenia, resulting in a high NLR. The increase in granulocytes is the result of the cytokine storm induced by the virus and is responsible for tissue damage (Bonetti et al., 2020)(Bohn et al., 2020).

A further remark concerning the blood analytes is that, in the BS-EWM, the thresholds of the single analytes exceeding the 0.5 death-risk closely overlap with the values recently proposed by other authors (Webb et al., 2020)(Caricchio et al., 2021) (Figure 2, panel B).

The present study is not unique in encompassing radiological findings combined with blood analysis. The study by Schalekamp et al. (Schalekamp et al., 2020) integrated blood analysis parameters and radiological information derived by grading chest X-rays (0–8 scale points). Unlike the cited study, with the BS-EWM in this study, the radiological score did not reach a high relevance (rel VIM) in predicting high risk. This difference can be explained by the different approaches used to build the model (Logistic regression vs Random Forests) and by the high degree of correlation of the X-ray score with multiple blood analytes: “collinearity” thus could have “stolen” importance from the information provided by imaging. Nevertheless, at admission, the chest X-ray score of patients who subsequently died was significantly higher than for patients who survived. Furthermore, the chest X-ray score may provide additional stability to the model, playing an important role in the case of missing data in the blood sample counterpart.

An important and pragmatic aspect offered by the BS-EWM is that the biomarkers employed may be obtained by the emergency laboratory in less than an hour (Garrafa et al., 2020a) and, differently from other biomarkers (Kyriazopoulou et al., 2021, p. 19), they are non-expensive and frequently used also in developing countries. It is important to note that the same methodology could be applied to other infections and be practical to triage people.

Most laboratories, including the small or peripheral ones, may provide results in a short time. At the Spedali Civili of Brescia, the BS-EWM is integrated within the Laboratory Information System. It works as a web-based calculator and is easy to interpret. It provides a risk threshold of 0.5, above which patients are graded as having a potentially high death-risk, thus supporting closer clinical observation or admission to a high-intensive care ward. In patients yielding a low risk (score 0 to 0.49), the decision by clinicians to allocate them to a low-intensive care ward or to monitoring is further sustained.

Finally, the need to regularly update models and closely monitor their performances over time and geographically should be underlined, given the rapidly changing nature of the disease and its management.

## Data Availability

The Institutional review board aprpoved the study with the entry code NP4000

## Supplementary Tables

**Table S.1:**
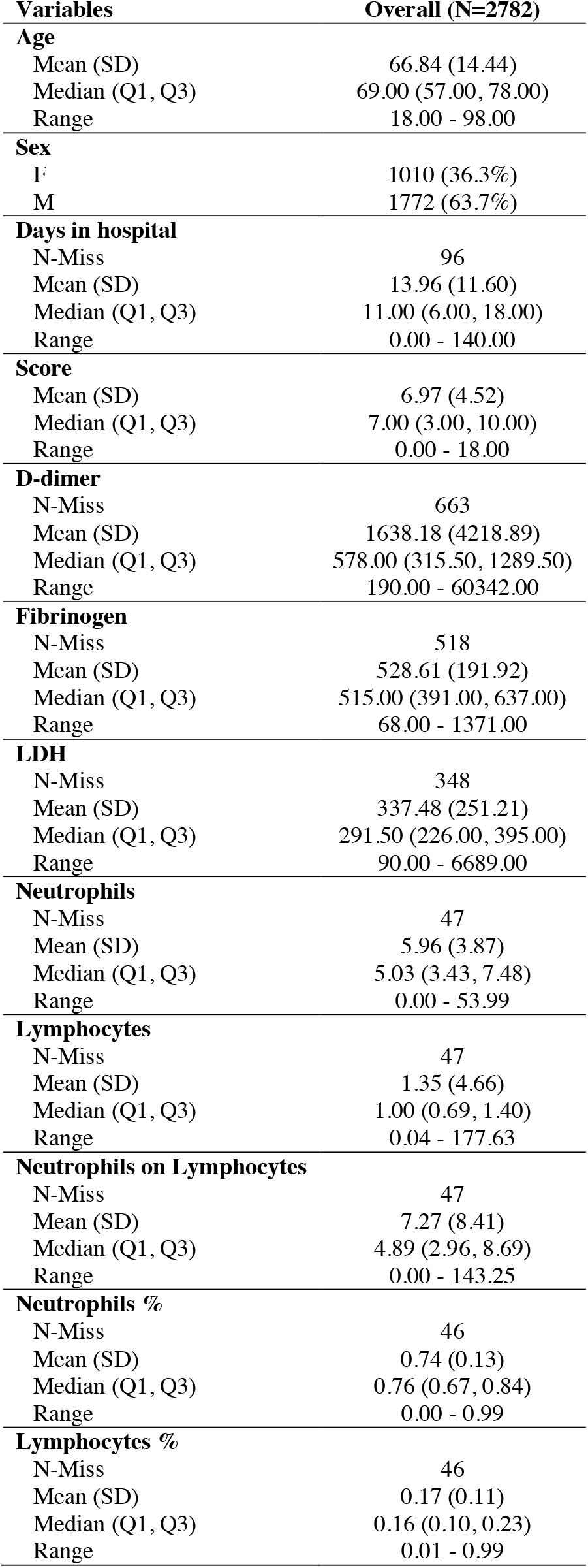
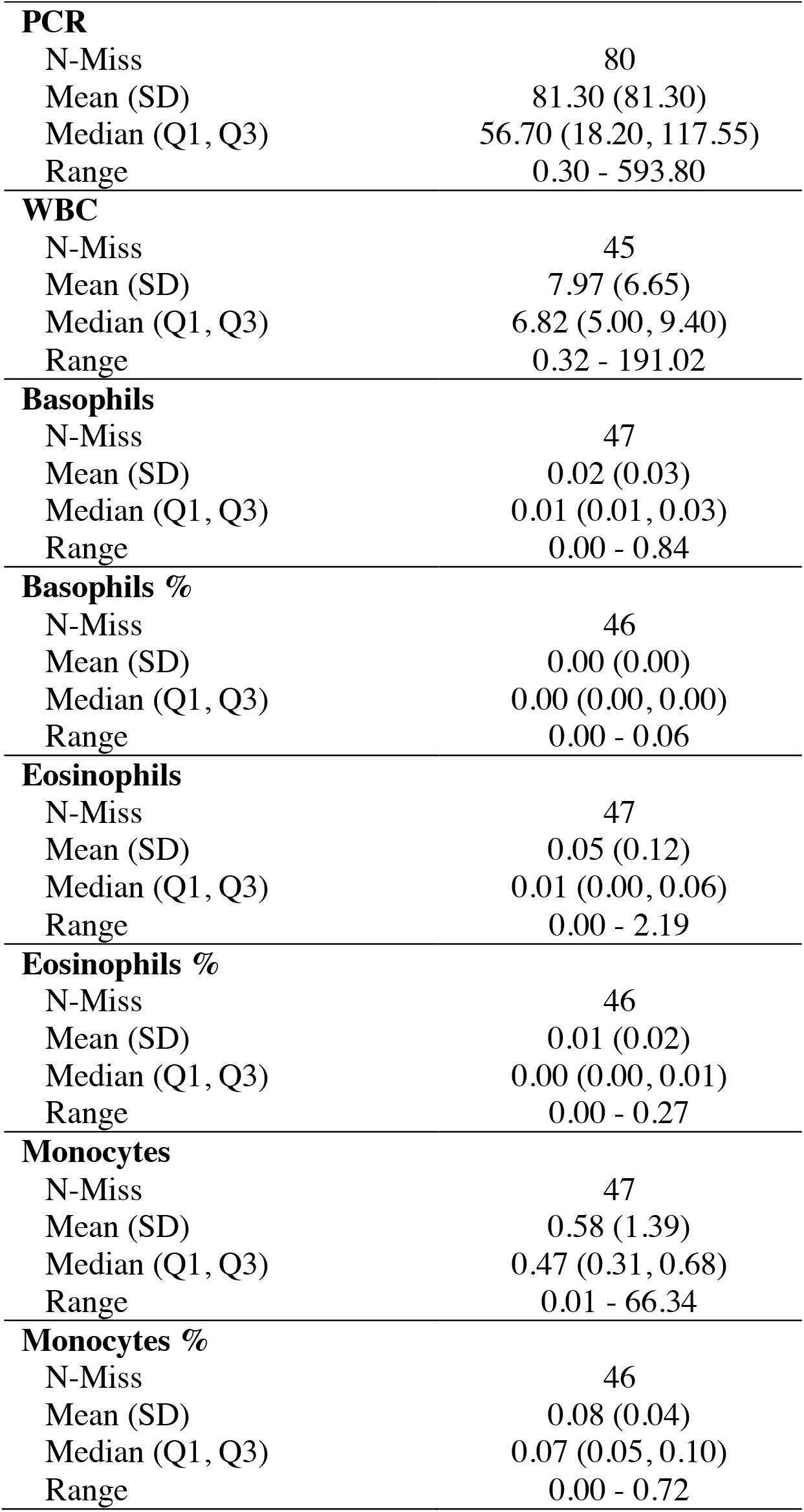
Descriptive statistics on all variables of the entire sample

**Table S.2:**
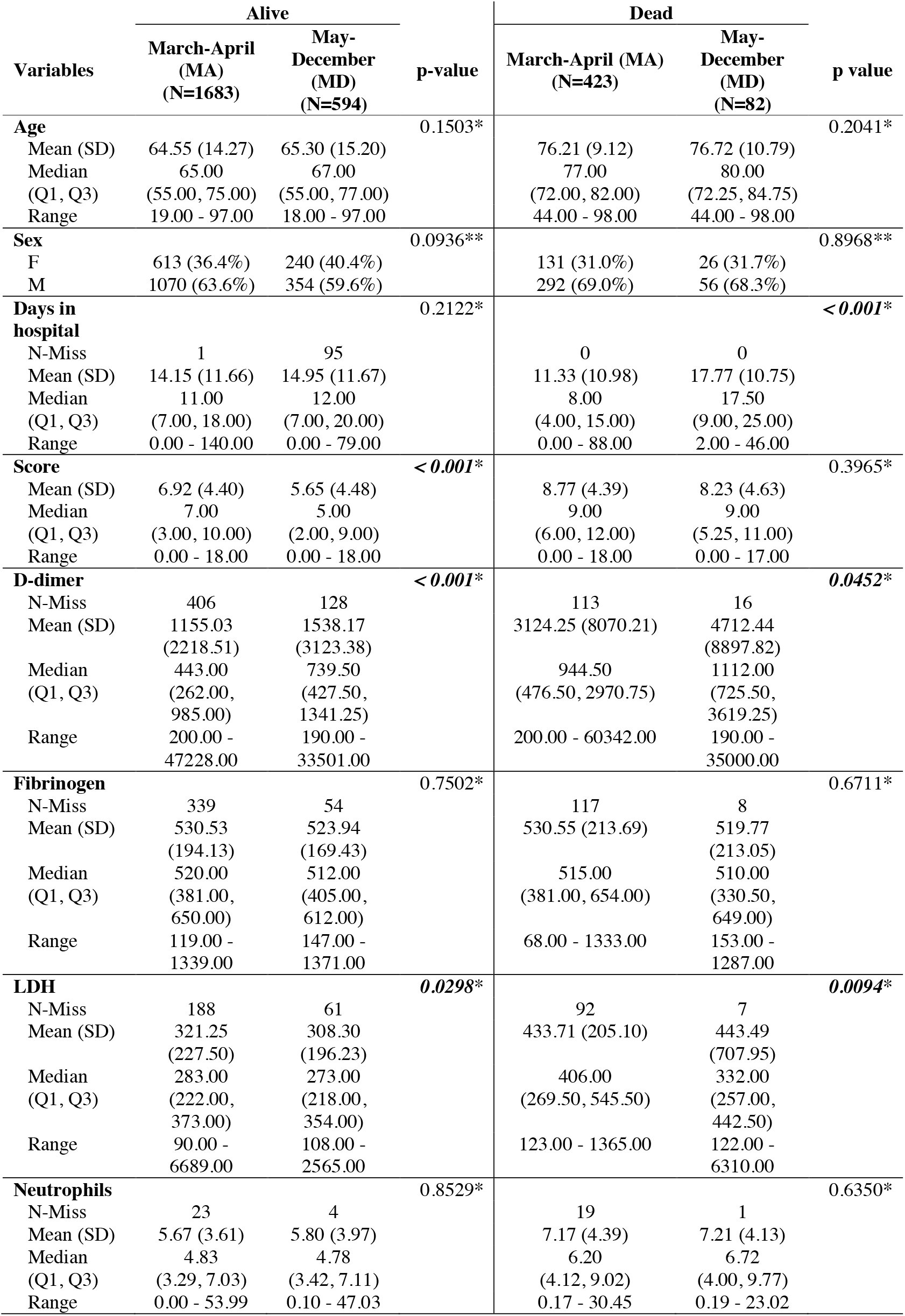
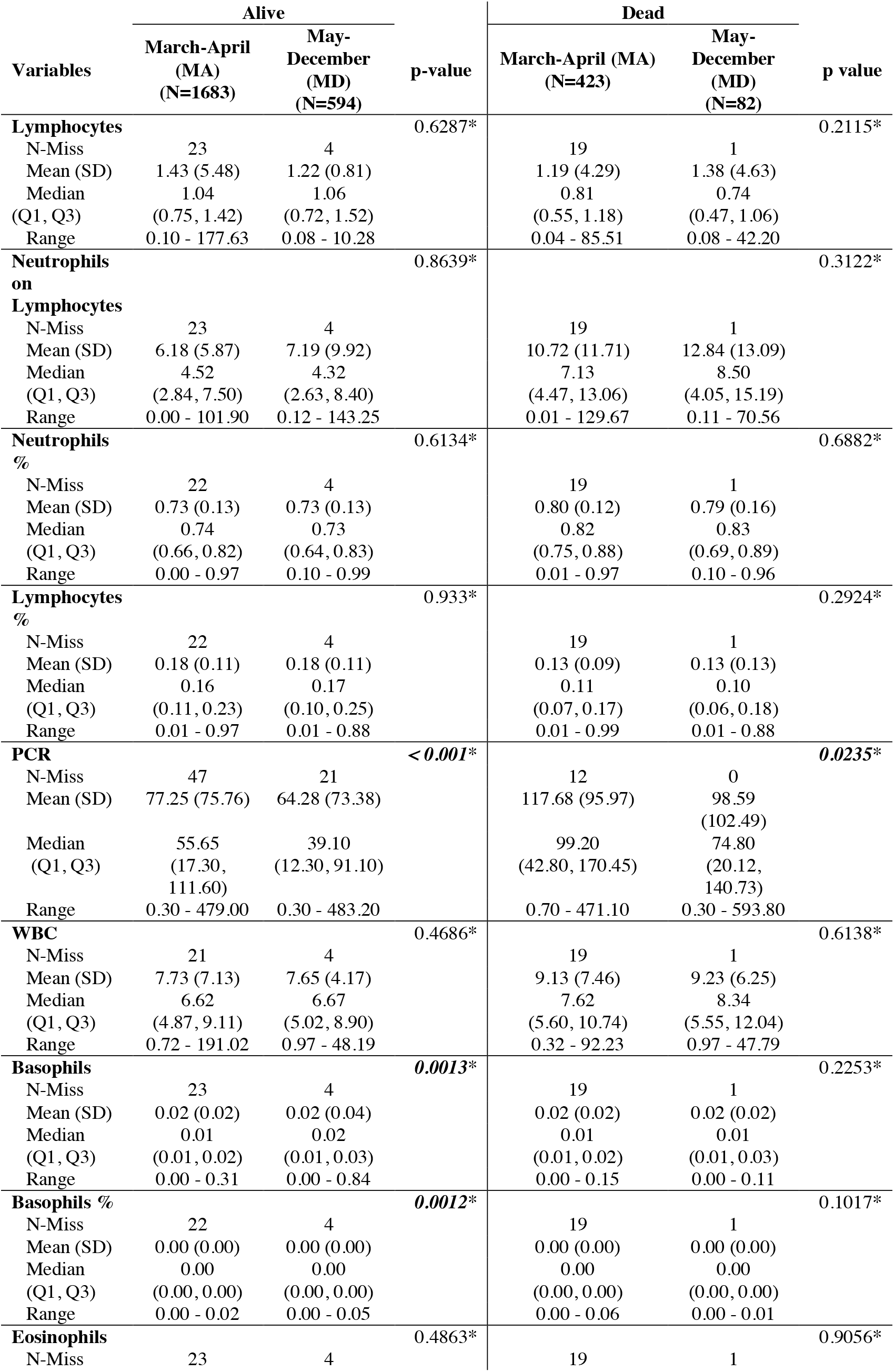
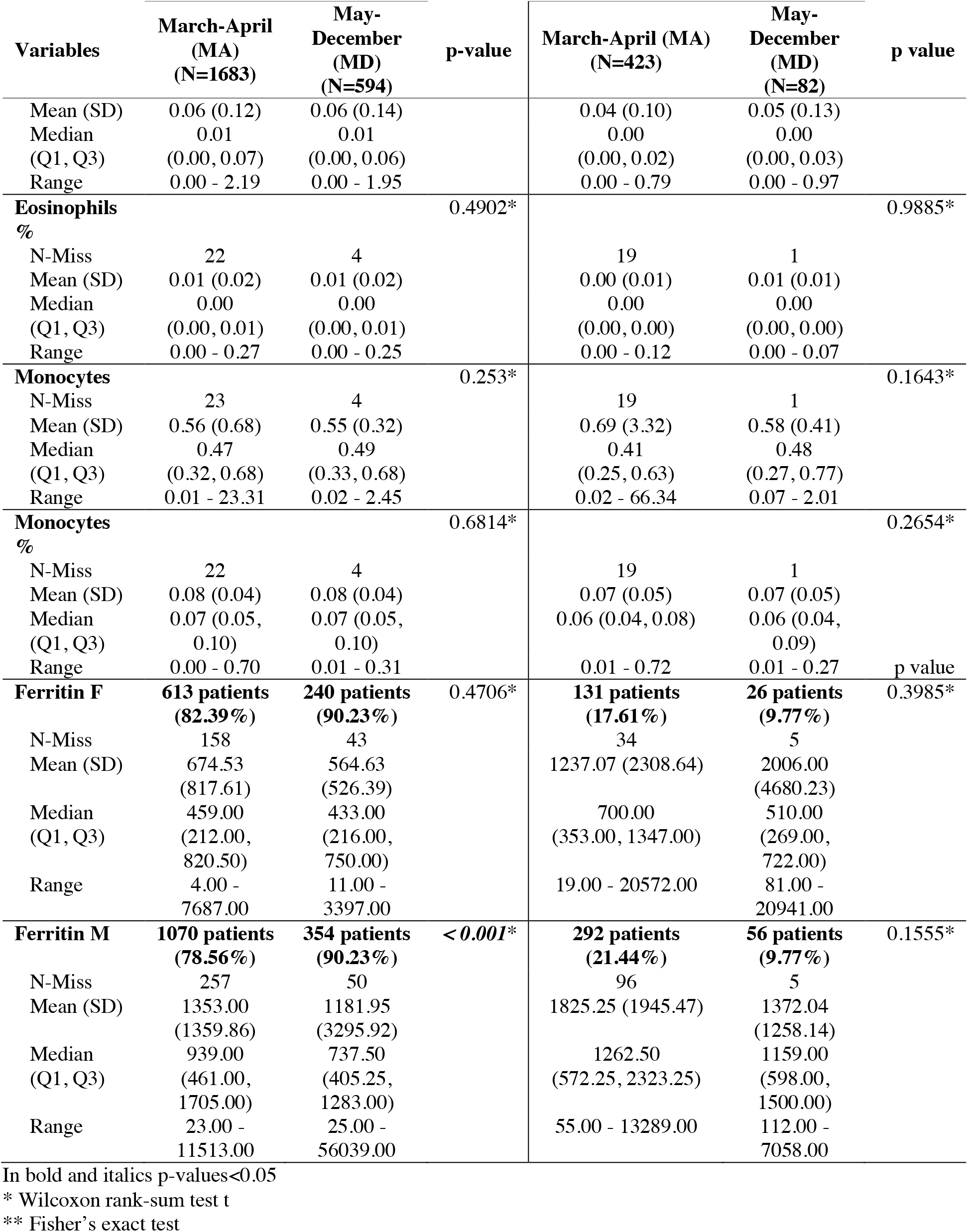
Descriptive statistics on all variables in the dataset stratified respect first (March-April 2020) and second (May-December 2020) wave. Comparison between Alive and Dead.

**Table S.3:** Performance metrics of the Random Forest (RF) using or not a rebalanced dataset with the SMOTE methodology

| Metrics | RF on a dataset rebalanced with the SMOTE methodology |  | RF on the original dataset |  |
| --- | --- | --- | --- | --- |
|  | Training<br>March-April (MA)<br>(95% CI) | Validating<br>March-April (MA)<br>(95% CI) | Training<br>March-April (MA)<br>(95% CI) | Validating<br>March-April (MA)<br>(95% CI) |
| **AUC (DeLong)** | 0.97-0.98 | 0.80-0.87 | 0.82-0.86 | 0.81-0.88 |
| **Sensitivity** | 0.93 (0.91-0.97) | 0.82 (0.72-0.92) | 0.81 (0.76-0.85) | 0.79 (0.66-0.95) |
| **Specificity** | 0.92 (0.88-0.94) | 0.75 (0.63-0.83) | 0.76 (0.73-0.79) | 0.76 (0.57-0.86) |

In this table we compare the performance of two RFs applied on (i) a dataset rebalanced with the SMOTE methodology (ii) the original dataset. This analysis suggests the use of SMOTE methodology before applying RF since the performance in Training and Validating groups (especially in terms of sensitivity) are better respect those obtained from the RF grown on the original dataset.

**Figure S.1:**
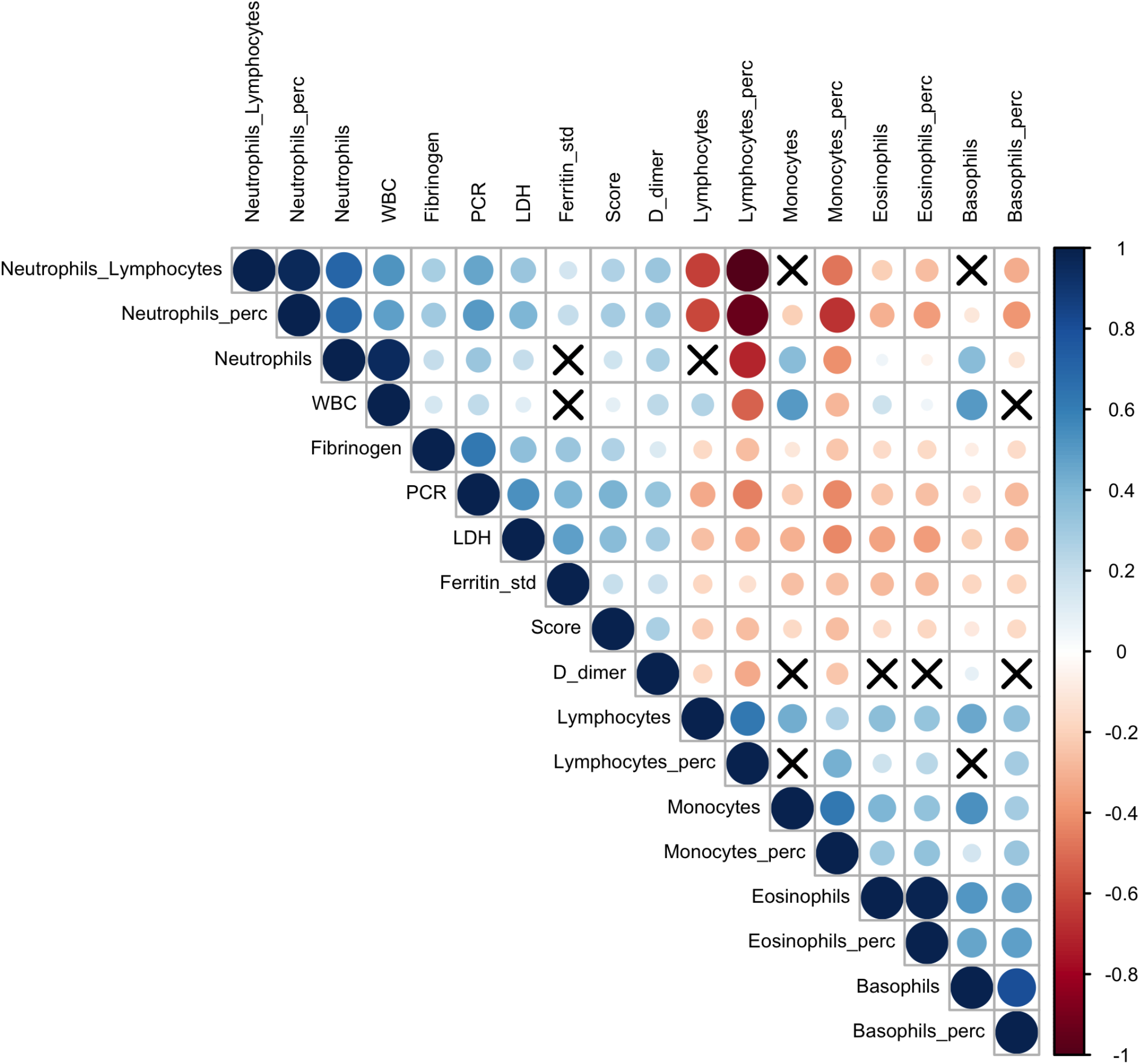
Correlation plot between the 17 analytes and Brescia chest-xray The relationships between 17 analytes and Brescia chest-xray score are inspected with the Spearman correlation coefficients, *ρ*_*s*_ which are represented in this correlation plot by means of blue and red circles (positive and negative correlation, respectively). The diameter of the circle is proportional to the magnitude of *ρ*_*s*_ and black crosses on them identify correlation not significantly different from zero (p-values>0.05). The correlation matrix is reordered according to the hierarchical cluster analysis on the quantitative variables.

## References

Anurag A, Preetam M. n.d. Validation of PSI/PORT, CURB-65 and SCAP scoring system in COVID-19 pneumonia for prediction of disease severity and 14-day mortality. Clin Respir J n/a. doi:https://doi.org/10.1111/crj.13326

Artero A, Madrazo M, Fernández-Garcés M, Muiño Miguez A, González García A, Crestelo Vieitez A, García Guijarro E, Fonseca Aizpuru EM, García Gómez M, Areses Manrique M, Martinez Cilleros C, Fidalgo Moreno MDP, Loureiro Amigo J, Gil Sánchez R, Rabadán Pejenaute E, Abella Vázquez L, Cañizares Navarro R, Solís Marquínez MN, Carrasco Sánchez FJ, González Moraleja J, Montero Rivas L, Escobar Sevilla J, Martín Escalante MD, Gómez-Huelgas R, Ramos-Rincón JM, SEMI-COVID-19 Network. 2021. Severity Scores in COVID-19 Pneumonia: a Multicenter, Retrospective, Cohort Study. J Gen Intern Med. doi:10.1007/s11606-021-06626-7

Avouac J, Drumez E, Hachulla E, Seror R, Georgin-Lavialle S, Mahou SE, Pertuiset E, Pham T, Marotte H, Servettaz A, Domont F, Chazerain P, Devaux M, Claudepierre P, Langlois V, Mekinian A, Maria ATJ, Banneville B, Fautrel B, Pouchot J, Thomas T, Flipo R-M, Richez C. 2021. COVID-19 outcomes in patients with inflammatory rheumatic and musculoskeletal diseases treated with rituximab: a cohort study. Lancet Rheumatol 0. doi:10.1016/S2665-9913(21)00059-X

Bohn MK, Lippi G, Horvath A, Sethi S, Koch D, Ferrari M, Wang C-B, Mancini N, Steele S, Adeli K. 2020. Molecular, serological, and biochemical diagnosis and monitoring of COVID-19: IFCC taskforce evaluation of the latest evidence. Clin Chem Lab Med 58:1037–1052. doi:10.1515/cclm-2020-0722

Bonetti G, Manelli F, Patroni A, Bettinardi A, Borrelli G, Fiordalisi G, Marino A, Menolfi A, Saggini S, Volpi R, Anesi A, Lippi G. 2020. Laboratory predictors of death from coronavirus disease 2019 (COVID-19) in the area of Valcamonica, Italy. Clin Chem Lab Med CCLM 58:1100–1105. doi:10.1515/cclm-2020-0459

Borghesi A, Maroldi R. 2020. COVID-19 outbreak in Italy: experimental chest X-ray scoring system for quantifying and monitoring disease progression. Radiol Med (Torino) 125:509–513. doi:10.1007/s11547-020-01200-3

Borghesi A, Zigliani A, Golemi S, Carapella N, Maculotti P, Farina D, Maroldi R. 2020a. Chest X-ray severity index as a predictor of in-hospital mortality in coronavirus disease 2019: A study of 302 patients from Italy. Int J Infect Dis 96:291–293. doi:10.1016/j.ijid.2020.05.021

Borghesi A, Zigliani A, Masciullo R, Golemi S, Maculotti P, Farina D, Maroldi R. 2020b. Radiographic severity index in COVID-19 pneumonia: relationship to age and sex in 783 Italian patients. Radiol Med (Torino) 1–4. doi:10.1007/s11547-020-01202-1

Borghi MO, Beltagy A, Garrafa E, Curreli D, Cecchini G, Bodio C, Grossi C, Blengino S, Tincani A, Franceschini F, Andreoli L, Lazzaroni MG, Piantoni S, Masneri S, Crisafulli F, Brugnoni D, Muiesan ML, Salvetti M, Parati G, Torresani E, Mahler M, Heilbron F, Pregnolato F, Pengo M, Tedesco F, Pozzi N, Meroni PL. 2020. Anti-Phospholipid Antibodies in COVID-19 Are Different From Those Detectable in the Anti-Phospholipid Syndrome. Front Immunol 11. doi:10.3389/fimmu.2020.584241

Breiman L. 2001. Random Forests. Mach Learn 45:5–32. doi:10.1023/A:1010933404324

Caricchio R, Gallucci M, Dass C, Zhang X, Gallucci S, Fleece D, Bromberg M, Criner GJ. 2021. Preliminary predictive criteria for COVID-19 cytokine storm. Ann Rheum Dis 80:88–95. doi:10.1136/annrheumdis-2020-218323

Carpita M, Vezzoli M. 2012. Statistical evidence of the subjective work quality: the fairness drivers of the job satisfaction. Electron J Appl Stat Anal 5:89–107. doi:10.1285/i20705948v5n1p89

Chawla NV, Bowyer KW, Hall LO, Kegelmeyer WP. 2002. SMOTE: Synthetic Minority Over-sampling Technique. J Artif Intell Res 16:321–357. doi:10.1613/jair.953

Codenotti S, Vezzoli M, Poliani PL, Cominelli M, Bono F, Kabbout H, Faggi F, Chiarelli N, Colombi M, Zanella I, Biasiotto G, Montanelli A, Caimi L, Monti E, Fanzani A. 2016. Caveolin-1, Caveolin-2 and Cavin-1 are strong predictors of adipogenic differentiation in human tumors and cell lines of liposarcoma. Eur J Cell Biol 95:252–264. doi:10.1016/j.ejcb.2016.04.005

Collins GS, Reitsma JB, Altman DG, Moons KGM. 2015. Transparent reporting of a multivariable prediction model for individual prognosis or diagnosis (TRIPOD): the TRIPOD statement. BMJ 350:g7594. doi:10.1136/bmj.g7594

Dancelli L, Manisera M, Vezzoli M. 2013. On Two Classes of Weighted Rank Correlation Measures Deriving from the Spearman’s ρ In: Giudici P, Ingrassia S, Vichi M, editors. Statistical Models for Data Analysis, Studies in Classification, Data Analysis, and Knowledge Organization. Heidelberg: Springer International Publishing. pp. 107–114. doi:10.1007/978-3-319-00032-9_13

Doglietto F, Vezzoli M, Biroli A, Saraceno G, Zanin L, Pertichetti M, Calza S, Agosti E, Aliaga Arias JM, Assietti R, Bellocchi S, Bernucci C, Bistazzoni S, Bongetta D, Fanti A, Fioravanti A, Fiorindi A, Franzin A, Locatelli D, Pugliese R, Roca E, Sicuri GM, Stefini R, Venturini M, Vivaldi O, Zattra C, Zoia C, Fontanella MM. 2020a. Anxiety in neurosurgical patients undergoing nonurgent surgery during the COVID-19 pandemic. Neurosurg Focus 49:E19. doi:10.3171/2020.9.FOCUS20681

Doglietto F, Vezzoli M, Gheza F, Lussardi GL, Domenicucci M, Vecchiarelli L, Zanin L, Saraceno G, Signorini L, Panciani PP, Castelli F, Maroldi R, Rasulo FA, Benvenuti MR, Portolani N, Bonardelli S, Milano G, Casiraghi A, Calza S, Fontanella MM. 2020b. Factors Associated With Surgical Mortality and Complications Among Patients With and Without Coronavirus Disease 2019 (COVID-19) in Italy. JAMA Surg. doi:10.1001/jamasurg.2020.2713

Friedman JH. 2001. Greedy function approximation: A gradient boosting machine. Ann Stat 29:1189–1232. doi:10.1214/aos/1013203451

Garrafa E, Brugnoni D, Barbaro M, Andreoli L, Focà E, Salvetti M, Castelli F, Franceschini F, Piva S, Muiesan ML, Latronico N, Levaggi R. 2020a. Laboratory considerations amidst the coronavirus disease 2019 outbreak: the Spedali Civili in Brescia experience. Bioanalysis 12:1223–1230. doi:10.4155/bio-2020-0109

Garrafa E, Levaggi R, Miniaci R, Paolillo C. 2020b. When fear backfires: Emergency department accesses during the Covid-19 pandemic. Health Policy Amst Neth 124:1333–1339. doi:10.1016/j.healthpol.2020.10.006

Gidari A, Socio GVD, Sabbatini S, Francisci D. 2020. Predictive value of National Early Warning Score 2 (NEWS2) for intensive care unit admission in patients with SARS-CoV-2 infection. Infect Dis 52:698–704. doi:10.1080/23744235.2020.1784457

Hong S, Lynn HS. 2020. Accuracy of random-forest-based imputation of missing data in the presence of non-normality, non-linearity, and interaction. BMC Med Res Methodol 20:199. doi:10.1186/s12874-020-01080-1

Kyriazopoulou E, Panagopoulos P, Metallidis S, Dalekos GN, Poulakou G, Gatselis N, Karakike E, Saridaki M, Loli G, Stefos A, Prasianaki D, Georgiadou S, Tsachouridou O, Petrakis V, Tsiakos K, Kosmidou M, Lygoura V, Dareioti M, Milionis H, Papanikolaou IC, Akinosoglou K, Myrodia D-M, Gravvani A, Stamou A, Gkavogianni T, Katrini K, Marantos T, Trontzas IP, Syrigos K, Chatzis L, Chatzis S, Vechlidis N, Avgoustou C, Chalvatzis S, Kyprianou M, van der Meer JW, Eugen-Olsen J, Netea MG, Giamarellos-Bourboulis EJ. 2021. An open label trial of anakinra to prevent respiratory failure in COVID-19. eLife 10. doi:10.7554/eLife.66125

Lazar Neto F, Marino LO, Torres A, Cilloniz C, Meirelles Marchini JF, Garcia de Alencar JC, Palomeque A, Albacar N, Brandão Neto RA, Souza HP, Ranzani OT, Bortolotto AL, Müller Veiga AD, Bellintani AP, Fantinatti BL, Nicolao BR, Caldeira BT, Umehara Juck CE, Bueno CG, Takamune DJ, Guidotte DV, D’Souza EA, Oliveira Silva EC, Brito Miyaguchi ET, Gomes da Silva EM, Santos Moreira EL, Fonseca e Silva FM, de Paula Maroni Escudeiro G, Travessini G, Costa GB, Tibucheski dos Santos H, Omori IH, Baptista JM, Afonso Nascimento JP, de Góes Campos L, Lima LT, Boscolo L, Adsuara Pandolfi MC, de Oliveira Silva M, Sanches MP, Saad Menezes MC, Gonçalves Cimatti De Calasans MM, Lima de Faria MF, Bezerra Martins NA, Albuquerque de Moura P, Araújo Simões PA, Luna RB, Nishiaka RK, Miléo RC, de Souza Abreu R, Toccoli RW, Monsalvarga TC, Brito Medeiros VM, Filippo Fernandes YS, Simões AL, Tavares AA, Carvalho de Alves Pereira C, Ribeiro DR, Dias de Francesco D, Emerenciano DL, Pires de Campos EM, Moreira FL, Bortoleto FM, Martinez G, Wiebelling da Silva G, Martins GB, Leite Fortes JC, Dias Barreto LG, Silva de Rosa ML, Ursoline do Nascimento M, Pisciolaro RF, Xavier RA, Barbosa de Souza SF, Lisboa Netto TA, Ribeiro S, Faria C, Rahhal H, Padrão E, Valente F, Padovan Chio YH, Gomez Gomez LM. 2021. Community-acquired pneumonia severity assessment tools in patients hospitalized with COVID-19: a validation and clinical applicability study. Clin Microbiol Infect. doi:10.1016/j.cmi.2021.03.002

Lazzaroni MG, Piantoni S, Masneri S, Garrafa E, Martini G, Tincani A, Andreoli L, Franceschini F. 2021. Coagulation dysfunction in COVID-19: The interplay between inflammation, viral infection and the coagulation system. Blood Rev 46:100745. doi:10.1016/j.blre.2020.100745

Liang W, Liang H, Ou L, Chen B, Chen A, Li C, Li Y, Guan W, Sang L, Lu J, Xu Y, Chen G, Guo H, Guo J, Chen Z, Zhao Y, Li S, Zhang N, Zhong N, He J, for the China Medical Treatment Expert Group for COVID-19. 2020. Development and Validation of a Clinical Risk Score to Predict the Occurrence of Critical Illness in Hospitalized Patients With COVID-19. JAMA Intern Med 180:1081. doi:10.1001/jamainternmed.2020.2033

Linssen J, Ermens A, Berrevoets M, Seghezzi M, Previtali G, van der Sar-van der Brugge S, Russcher H, Verbon A, Gillis J, Riedl J, de Jongh E, Saker J, Münster M, Munnix IC, Dofferhof A, Scharnhorst V, Ammerlaan H, Deiteren K, Bakker SJ, Van Pelt LJ, Kluiters-de Hingh Y, Leers MP, van der Ven AJ. 2020. A novel haemocytometric COVID-19 prognostic score developed and validated in an observational multicentre European hospital-based study. eLife 9:e63195. doi:10.7554/eLife.63195

Lippi G, Henry BM, Hoehn J, Benoit S, Benoit J. 2020. Validation of the Corona-Score for rapid identification of SARS-CoV-2 infections in patients seeking emergency department care in the United States. Clin Chem Lab Med CCLM 58:e311–e313. doi:10.1515/cclm-2020-1121

Marengoni A, Zucchelli A, Vetrano DL, Armellini A, Botteri E, Nicosia F, Romanelli G, Beindorf EA, Giansiracusa P, Garrafa E, Ferrucci L, Fratiglioni L, Bernabei R, Onder G. 2021. Beyond Chronological Age: Frailty and Multimorbidity Predict In-Hospital Mortality in Patients With Coronavirus Disease 2019. J Gerontol Ser A 76:e38–e45. doi:10.1093/gerona/glaa291

Maroldi R, Rondi P, Agazzi GM, Ravanelli M, Borghesi A, Farina D. 2020. Which role for chest x-ray score in predicting the outcome in COVID-19 pneumonia? Eur Radiol. doi:10.1007/s00330-020-07504-2

Martens RJH, Adrichem AJ van, Mattheij NJA, Brouwer CG, Twist DJL van, Broerse JJCR, Magro-Checa C, Dongen CMP van, Mostard RLM, Ramiro S, Landewé RBM, Leers MPG. 2021. Hemocytometric characteristics of COVID-19 patients with and without cytokine storm syndrome on the sysmex XN-10 hematology analyzer. Clin Chem Lab Med CCLM 59:783–793. doi:10.1515/cclm-2020-1529

Marziano M, Tonello S, Cantù E, Abate G, Vezzoli M, Rungratanawanich W, Serpelloni M, Lopomo NF, Memo M, Sardini E, Uberti D. 2019. Monitoring Caco-2 to enterocyte-like cells differentiation by means of electric impedance analysis on printed sensors. Biochim Biophys Acta Gen Subj 1863:893–902. doi:10.1016/j.bbagen.2019.02.008

Myrstad M, Ihle-Hansen H, Tveita AA, Andersen EL, Nygård S, Tveit A, Berge T. 2020. National Early Warning Score 2 (NEWS2) on admission predicts severe disease and in-hospital mortality from Covid-19 – a prospective cohort study. Scand J Trauma Resusc Emerg Med 28:66. doi:10.1186/s13049-020-00764-3

Salvi A, Vezzoli M, Busatto S, Paolini L, Faranda T, Abeni E, Caracausi M, Antonaros F, Piovesan A, Locatelli C, Cocchi G, Alvisi G, De Petro G, Ricotta D, Bergese P, Radeghieri A. 2019. Erratum: Analysis of a nanoparticle-enriched fraction of plasma reveals miRNA candidates for down syndrome pathogenesis(Int J Mol Med (2019) 43(2303-2318) DOI: 10.3892/ijmm.2019.4158). Int J Mol Med 44:768. doi:10.3892/ijmm.2019.4222

Schalekamp S, Huisman M, van Dijk RA, Boomsma MF, Freire Jorge PJ, de Boer WS, Herder GJM, Bonarius M, Groot OA, Jong E, Schreuder A, Schaefer-Prokop CM. 2020. Model-based Prediction of Critical Illness in Hospitalized Patients with COVID-19. Radiology 298:E46–E54. doi:10.1148/radiol.2020202723

Sperrin M, Grant SW, Peek N. 2020. Prediction models for diagnosis and prognosis in Covid-19. BMJ 369:m1464. doi:10.1136/bmj.m1464

Vezzoli M. 2011. Exploring the facets of overall job satisfaction through a novel ensemble learning. Electron J Appl Stat Anal 4:23-38–38. doi:10.1285/i20705948v4n1p23

Vezzoli M, Ravaggi A, Zanotti L, Miscioscia RA, Bignotti E, Ragnoli M, Gambino A, Ruggeri G, Calza S, Sartori E, Odicino F. 2017. RERT: A Novel Regression Tree Approach to Predict Extrauterine Disease in Endometrial Carcinoma Patients. Sci Rep 7:10528. doi:10.1038/s41598-017-11104-4

Webb BJ, Peltan ID, Jensen P, Hoda D, Hunter B, Silver A, Starr N, Buckel W, Grisel N, Hummel E, Snow G, Morris D, Stenehjem E, Srivastava R, Brown SM. 2020. Clinical criteria for COVID-19-associated hyperinflammatory syndrome: a cohort study. Lancet Rheumatol 2:e754–e763. doi:10.1016/S2665-9913(20)30343-X

Wynants L, Calster BV, Collins GS, Riley RD, Heinze G, Schuit E, Bonten MMJ, Dahly DL, Damen JA, Debray TPA, Jong VMT de, Vos MD, Dhiman P, Haller MC, Harhay MO, Henckaerts L, Heus P, Kammer M, Kreuzberger N, Lohmann A, Luijken K, Ma J, Martin GP, McLernon DJ, Navarro CLA, Reitsma JB, Sergeant JC, Shi C, Skoetz N, Smits LJM, Snell KIE, Sperrin M, Spijker R, Steyerberg EW, Takada T, Tzoulaki I, Kuijk SMJ van, Bussel BCT van, Horst ICC van der, Royen FS van, Verbakel JY, Wallisch C, Wilkinson J, Wolff R, Hooft L, Moons KGM, Smeden M van. 2020. Prediction models for diagnosis and prognosis of covid-19: systematic review and critical appraisal. BMJ 369:m1328. doi:10.1136/bmj.m1328

Yavuz BG, Colak S, Guven R, Altundag İ, Seyhan AU, Inanc RG. 2021. Clinical Features of the 60 Years and Older Patients Infected with 2019 Novel Coronavirus: Can We Predict Mortality Earlier? Gerontology 1–8. doi:10.1159/000514481

